# Results of an Academic Dialysis Program-Wide SARS-CoV-2 Vaccination Effort

**DOI:** 10.1101/2021.05.07.21256841

**Authors:** Brendan T. Bowman, Benjamin J. Lobo, Binu Sharma, Jennie Z. Ma

## Abstract

Patients with End Stage Kidney Disease requiring dialysis are exceedingly vulnerable to SARS-CoV-2 infection with high hospitalization rates and mortality. Despite this risk, little is known about real world dialysis patient SARS-CoV-2 vaccination acceptance. Surveys of the general population suggest significant vaccine hesitancy and high potential for refusal. From January 27^th^ to March 12^th^ 2021, the University of Virginia (UVA) Health System, in partnership with the Virginia Department of Health / Blue Ridge Health District (BRHD) provided on-site mobile vaccination clinics at 12 UVA dialysis sites. We conducted a cross-sectional study to evaluate vaccine acceptance and evaluate factors associated with refusal. Of 859 dialysis patients with complete vaccination data, 80% received at least one dose of vaccine and 87% of these vaccinations were provided by the UVA/BRHD partnership. The overall patient refusal rate was low at 14%. Patients refusing SARS-CoV-2 vaccine were more likely to be female, younger and missing a documented flu vaccination during the 2020-2021 season. Attributes such as race or prior infection with SARS-CoV-2 were not significantly associated with vaccine refusal. In conclusion, dialysis patients in our program were surprisingly likely to accept vaccination for SARS-CoV-2. Identifying attributes associated with refusal may help target populations at high risk of vaccine refusal.

## Introduction

Chronic kidney disease (CKD) is a significant risk factor for poor health outcomes following infection with SARS-CoV-2 (1). The most severe form of CKD, End Stage Kidney Disease (ESKD), has particularly poor health outcomes with infection related mortality rates ranging from 20-30% in retrospective reviews (2). Given these poor outcomes, there have been strong advocacy efforts to obtain broad vaccine access for patients with ESKD (3). Despite these efforts, dialysis dependent patients were mostly excluded from the earliest “1a” wave of vaccinations leading to delays in real world assessment of vaccine administration efforts. Only recently has vaccine become available at scale to dialysis patients and providers (4) As such, there is a dearth of real world data on vaccine acceptance rates among U.S. dialysis patients. National surveys of the general public obtained early in the period suggested vaccine acceptance may range as low as 58% to 69% (5, 6). In these studies, various factors were found to be associated with vaccine hesitancy including: younger age, African American race, lower attained education level and lower influenza vaccination rates (5). A more recent national survey of dialysis patients suggested vaccine acceptance rates could reach as high as 81% under ideal conditions (7). In that survey, similar factors were associated with vaccine hesitancy in the dialysis population as seen in the general population.

In partnership with the Virginia Department of Health’s Blue Ridge Health District (BRHD), the University of Virginia (UVA) Health System undertook one a dialysis program-wide vaccination effort from January to March of 2021. The campaign employed a series of mobile pharmacist and nurse led vaccination clinics visiting each UVA dialysis clinic. Here, we present the results and analysis of that initiative in respect to vaccine acceptance, refusal and an evaluation of factors associated with vaccine refusal.

## Methods

### Dialysis Vaccination Program

The University of Virginia Health System operates 12 dialysis clinics throughout central Virginia with associated peritoneal and home dialysis clinics co-located at select sites. UVA Health mobile vaccination clinics provided each eligible patient a two-shot series of the Pfizer-BioNTech COVID-19 vaccine (BNT162b2) over a series of 4 visits to each dialysis clinic. Peritoneal dialysis (PD) and home hemodialysis (HHD) patients were offered vaccine on the same days and intervals as in-center hemodialysis patients. Vaccinations were administered by dialysis pharmacists, nurses and advanced practice nurses. Vaccine educational information was provided to patients on the day of vaccination and consent or refusal obtained and documented during the chairside visit.

### Data Collection

Included in the final analysis were dialysis patients ≥ 18 years of age with ESKD registered to a UVA facility on the first day of the mobile vaccination clinic for each respective dialysis clinic. Patients dialyzing with acute kidney injury, transient patients, and those registering at a unit after the vaccination effort began, were excluded, yielding a total of 872 vaccine-eligible patients. Thirteen patients had incomplete data and were excluded, yielding a total of 859 patients for final analysis (Figure 1). To analyze risk factors associated with vaccine acceptance or refusal, demographic and clinical information was captured from the dialysis-specific electronic medical record system. Two proxy measures for socio-economic factors were used based on patient ZIP Code: median household income taken from Table B19013 of the 2019 American Community Survey 5-Year Estimates, and the mean Area Deprivation Index (ADI) national percentile (8). Prior SARS-CoV-2 infection was captured from a program-wide COVID-19 tracking tool. Data collection concluded on March 16^th^, 2021.

**Figure 1:**
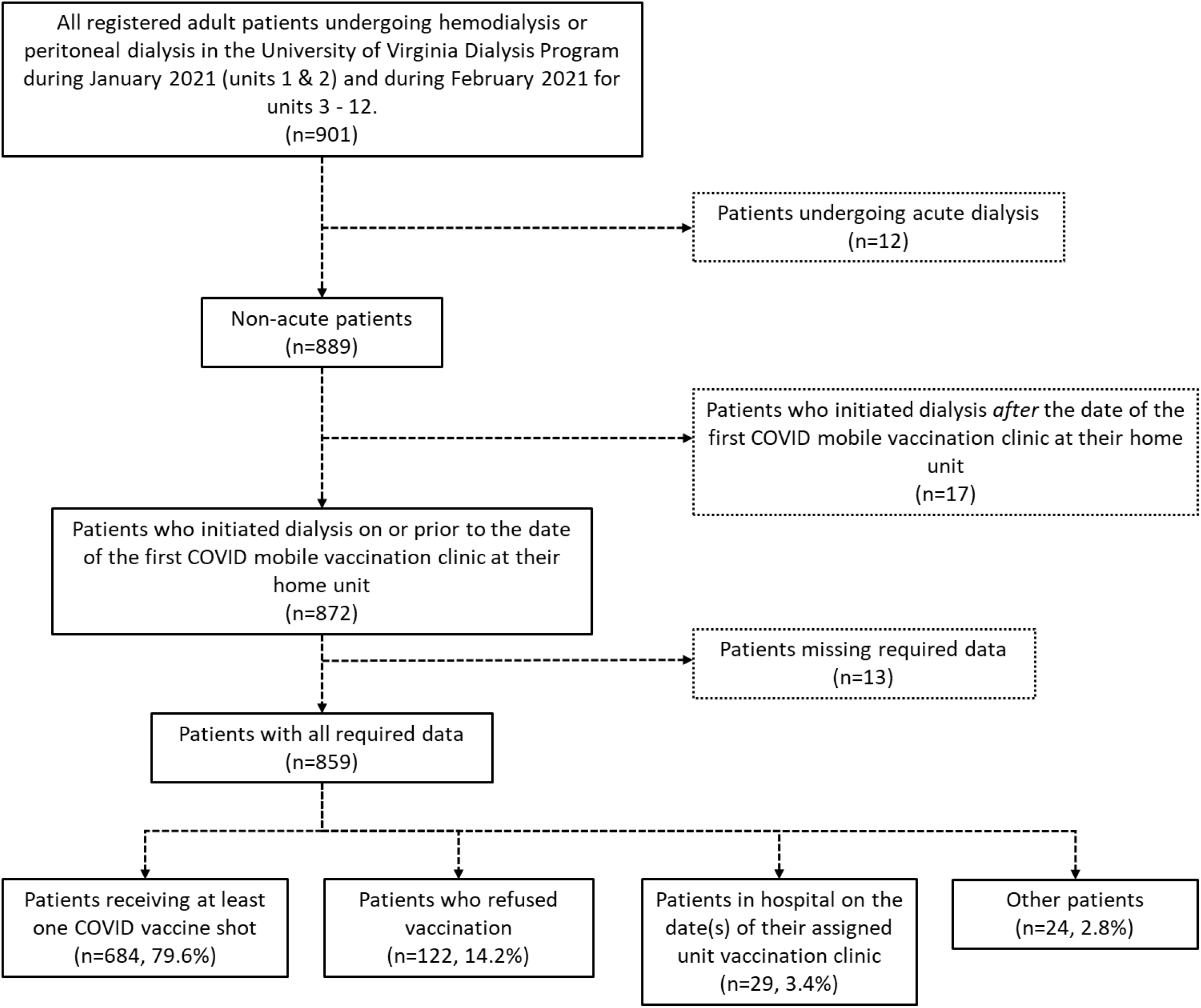
Patient Inclusion Flow Chart

Based on COVID vaccination data, patients were classified into 4 groups: Vaccinated, Refused, Hospitalized and Other. The ‘Vaccinated’ group was defined as patients that received at least one shot at either an on-site UVA/BRHD mobile clinic or an offsite provider/clinic; the ‘Refused’ group was defined as vaccine eligible patients present in the dialysis clinic at the time of the mobile vaccination clinic who declined offered vaccination; the ‘Hospitalized’ group was defined as those patients who were inpatient on the date(s) of initial COVID vaccination clinic at their home units (unable to receive vaccination); the ‘Other’ group included all remaining patients; the vast majority of this group constituted patients who were not present at their dialysis clinics on the dates of the COVID vaccination clinic (“no-shows”). Due to limited vaccine supply, catch up clinics were not offered for those unable to receive a first shot.

In addition to patient-level data, dialysis clinic level data was collected including infection rate and census. The infection rate for each dialysis clinic was calculated as a function of the number of infections per patient exposure months from Nov 1, 2020 through Feb 28, 2021, roughly correlating with the second surge in central Virginia. Unit census was determined as the number of patients registered at the facility on the date of the first on-site vaccination clinic.

### Statistical Analysis

All 859 dialysis patients were included in the descriptive analysis of patient characteristics and risk factors, but only those in the ‘Vaccinated’ group (N=684) and those in the ‘Refused’ group (N=122) were included in the subsequent statistical testing and regression analyses. Data were summarized as mean and standard deviation or median (25^th^, 75^th^ percentiles) for continuous variables and as frequency and percentage for categorical variables. The difference in patient characteristics between the ‘Vaccinated’ and ‘Refused’ groups were evaluated using Fisher’s exact test or Chi-square test for categorical variables and parametric t-test or Mann-Whitney test for continuous variables.

The primary response of interest in this study was vaccine acceptance or refusal within the study window. We modeled the probability of vaccine refusal using a logistic regression model to evaluate the effects of patient characteristics and risk factors on the outcome. Prior to the regression analyses, we first estimated the intra-class correlation (ICC) to assess the clustering effect in dialysis facility units and found a small yet statistically significant ICC=0.05. To account for this clustering effect in the dialysis units, the probability of vaccine refusal was modeled in logistic regression with the generalized estimating equation (GEE) method. The patient characteristics and risk factors were initially evaluated in a univariate fashion. Continuous variables were carefully assessed for their relationship with the probability of refusal on the logit scale, and those variables with non-linear relationship were further categorized. The final multivariable model was derived using stepwise selection in GEE with the minimum quasi-information criterion (the pstools package in R, https://rdrr.io/github/dewittpe/pstools/src/R/gee_stepper.R). The effects of patient characteristics and risk factors were measured and reported by odds ratios (OR) of vaccine refusal and their corresponding 95% confidence interval (CI) along with the *p*-values. A *p*-value < 0.05 was considered to be statistically significant. All data analyses were performed using R software version 3.6.3 (http://www.R-project.org/). The research proposal was evaluated by the UVA Institutional Review Board for Health Sciences Research and found to be exempt from IRB review / not human subjects research (tracking #23147)

## Results

Within the UVA dialysis program, a total of 872 patients were eligible to receive a SARS-CoV-2 vaccine, with 859 patients included in the final analysis. Overall, the average age was 64 ± 14 years, 44.7% were females, 51% were black, and 88.5% were on in-center hemodialysis compared to 11.5% home dialysis (PD or HHD). The complete demographic and clinical characteristics of all patients are reported in Table 1.

**Table 1:**
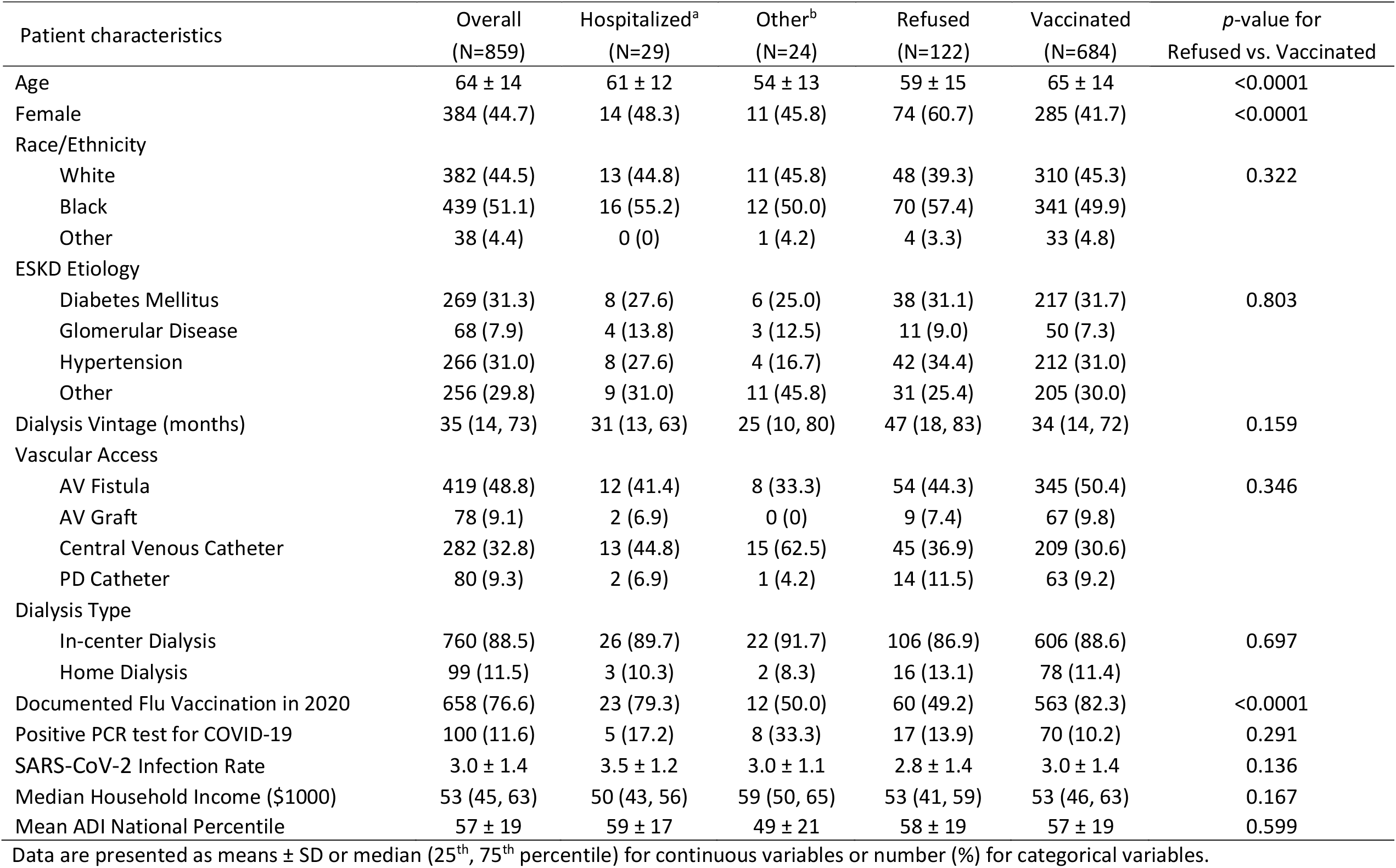
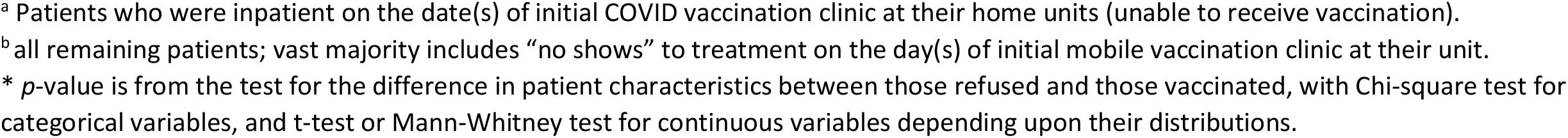
Descriptive summary of patient characteristics in the COVID-19 Vaccination Study

| Patient characteristics | Overall<br>(N=859) | Hospitalized<sup>a</sup><br>(N=29) | Other<sup>b</sup><br>(N=24) | Refused<br>(N=122) | Vaccinated<br>(N=684) | *p*-value for<br>Refused vs. Vaccinated |
| --- | --- | --- | --- | --- | --- | --- |
| Age | 64 ± 14 | 61 ± 12 | 54 ± 13 | 59 ± 15 | 65 ± 14 | <0.0001 |
| Female | 384 (44.7) | 14 (48.3) | 11 (45.8) | 74 (60.7) | 285 (41.7) | <0.0001 |
| Race/Ethnicity |  |  |  |  |  |  |
| White | 382 (44.5) | 13 (44.8) | 11 (45.8) | 48 (39.3) | 310 (45.3) | 0.322 |
| Black | 439 (51.1) | 16 (55.2) | 12 (50.0) | 70 (57.4) | 341 (49.9) |  |
| Other | 38 (4.4) | 0 (0) | 1 (4.2) | 4 (3.3) | 33 (4.8) |  |
| ESKD Etiology |  |  |  |  |  |  |
| Diabetes Mellitus | 269 (31.3) | 8 (27.6) | 6 (25.0) | 38 (31.1) | 217 (31.7) | 0.803 |
| Glomerular Disease | 68 (7.9) | 4 (13.8) | 3 (12.5) | 11 (9.0) | 50 (7.3) |  |
| Hypertension | 266 (31.0) | 8 (27.6) | 4 (16.7) | 42 (34.4) | 212 (31.0) |  |
| Other | 256 (29.8) | 9 (31.0) | 11 (45.8) | 31 (25.4) | 205 (30.0) |  |
| Dialysis Vintage (months) | 35 (14, 73) | 31 (13, 63) | 25 (10, 80) | 47 (18, 83) | 34 (14, 72) | 0.159 |
| Vascular Access |  |  |  |  |  |  |
| AV Fistula | 419 (48.8) | 12 (41.4) | 8 (33.3) | 54 (44.3) | 345 (50.4) | 0.346 |
| AV Graft | 78 (9.1) | 2 (6.9) | 0 (0) | 9 (7.4) | 67 (9.8) |  |
| Central Venous Catheter | 282 (32.8) | 13 (44.8) | 15 (62.5) | 45 (36.9) | 209 (30.6) |  |
| PD Catheter | 80 (9.3) | 2 (6.9) | 1 (4.2) | 14 (11.5) | 63 (9.2) |  |
| Dialysis Type |  |  |  |  |  |  |
| In-center Dialysis | 760 (88.5) | 26 (89.7) | 22 (91.7) | 106 (86.9) | 606 (88.6) | 0.697 |
| Home Dialysis | 99 (11.5) | 3 (10.3) | 2 (8.3) | 16 (13.1) | 78 (11.4) |  |
| Documented Flu Vaccination in 2020 | 658 (76.6) | 23 (79.3) | 12 (50.0) | 60 (49.2) | 563 (82.3) | <0.0001 |
| Positive PCR test for COVID-19 | 100 (11.6) | 5 (17.2) | 8 (33.3) | 17 (13.9) | 70 (10.2) | 0.291 |
| SARS-CoV-2 Infection Rate | 3.0 ± 1.4 | 3.5 ± 1.2 | 3.0 ± 1.1 | 2.8 ± 1.4 | 3.0 ± 1.4 | 0.136 |
| Median Household Income (\$1000) | 53 (45, 63) | 50 (43, 56) | 59 (50, 65) | 53 (41, 59) | 53 (46, 63) | 0.167 |
| Mean ADI National Percentile | 57 ± 19 | 59 ± 17 | 49 ± 21 | 58 ± 19 | 57 ± 19 | 0.599 |

Data are presented as means ± SD or median (25<sup>th</sup>, 75<sup>th</sup> percentile) for continuous variables or number (%) for categorical variables.
<sup>a</sup> Patients who were inpatient on the date(s) of initial COVID vaccination clinic at their home units (unable to receive vaccination).
<sup>b</sup> all remaining patients; vast majority includes “no shows” to treatment on the day(s) of initial mobile vaccination clinic at their unit.
\* $p$ -value is from the test for the difference in patient characteristics between those refused and those vaccinated, with Chi-square test for categorical variables, and t-test or Mann-Whitney test for continuous variables depending upon their distributions.

**Table 2:**
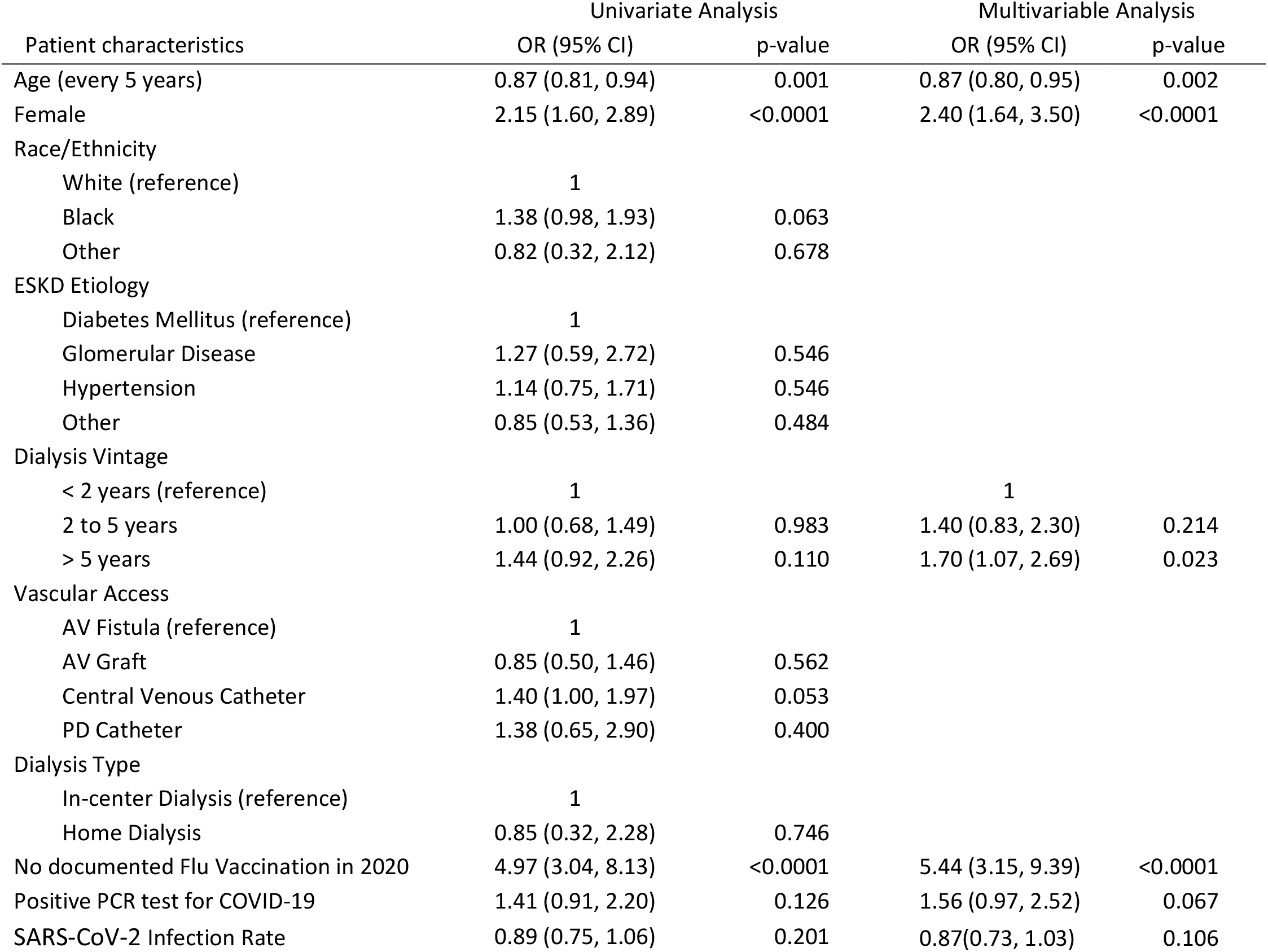
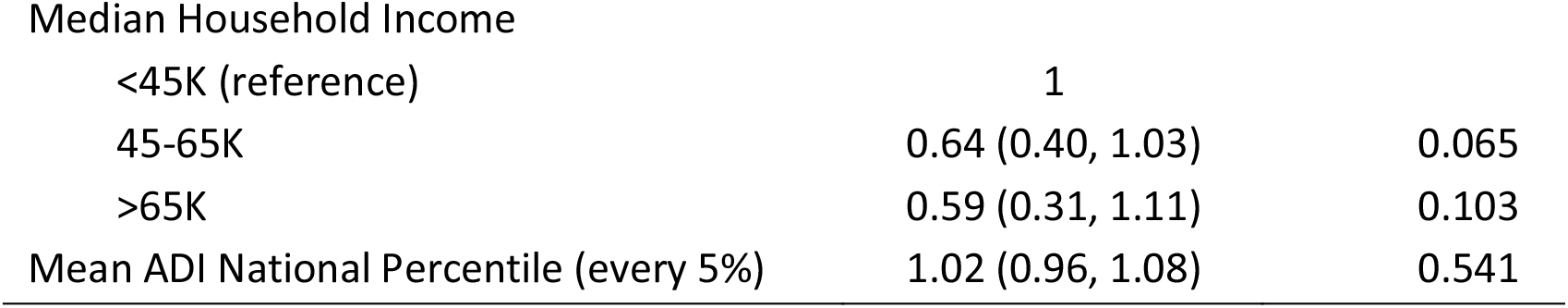
Patient characteristics associated with the likelihood of refusal in the COVID-19 Vaccination Study

Between the UVA/BRHD vaccination partnership and outside organizations, 684 (79.6%) of all eligible patients received at least one vaccination as of March 16^th^, 2021. Of these, 639 patients received the two shot series, including 58 skilled nursing facility residents vaccinated in the state’s 1a wave (45 patients missing data on 2^nd^ shot). Refusals occurred in 14.2% of all patients. The vast majority of the remaining 6.2% remaining unvaccinated were either due to hospitalization or no-shows to treatment on the vaccine clinic day. Excluding the 6.2% of patients that were absent or hospitalized, 85% of the remaining 806 dialysis patients who received an in-person offer of vaccination were vaccinated. Of these 806 patients, those refusing SARS-CoV-2 vaccination (n=122, 15%) were younger, more likely to be female and more likely to lack a documented influenza vaccination in 2020 (Table 1).

To further characterize variables associated with vaccine refusal a univariate and multivariable analysis with GEE was undertaken. In the univariate analysis, comparison of the ‘Vaccinated’ versus ‘Refused’ groups shows they were similar in terms of home versus in-center distribution and clinical characteristics such as ESKD etiology, and dialysis vintage. Categorized dialysis vintage and median household income in the ZIP Code where patients reside were considered for their nonlinear relationship with the probability of refusal in logit scale. Age, gender and absent documentation of a flu shot in 2020 were significantly associated with the likelihood of vaccine refusal. Black race/ethnicity, central venous catheter use and residence in a Zip code with a median income between $45K and $65K were only marginally associated with the outcome (*p*=0.063, *p*=0.053, and *p*=0.065 respectively). The local dialysis clinic SARS-CoV-2 infection rate over the preceding four-month period (November 1 2020 to February 28, 2021, corresponding to the local COVID-19 peak surge period) was lower in the ‘Refused’ group though not statistically significant. Likewise, the proportion of PCR-proven SARS-CoV-2 infection was slightly higher in the ‘Refused’ group (13.9% vs. 10.2%), but this also was not significant.

Based on the stepwise selection, the final multivariable model included age, gender, dialysis vintage, documentation of flu shot, SARS-CoV-2 PCR test result and infection rate. After multivariable adjustment, age, female sex, and lack of documented influenza vaccination for 2020 remained statistically significant. Specifically, the likelihood of refusal decreased by 13% for every 5-year increment in age (OR=0.87, 95% CI: 0.80 to 0.95, *p*=0.002). Women were more than twice as likely to refuse the vaccination as men (OR=2.40, 95% CI: 1.64 to 3.50, *p*<0.0001). Those without documentation of a flu shot in 2020 were more than five times as likely to refuse compared to those who had the flu shot in 2020 (OR=5.443, 95% CI: 3.15 to 9.39, *p*<0.0001). Interestingly, multivariable adjustment attenuated the effects of race, central venous catheter access type, and median household income, which were not selected in the final model, and amplified the effects of dialysis vintage and PCR test result for COVID-19, which became statistically or marginally significant. Notably, those on dialysis for more than 5 years were 1.7 times more likely to refuse vaccination (OR=1.7, 95% CI: 1.08 to 2.70, *p*=0.023), and those with a positive PCR test for COVID-19 were 1.58 times more likely to refuse vaccination (OR=1.56, 95% CI: 0.97 to 2.52, *p*=0.067). The local dialysis clinic SARS-CoV-2 infection rate was included in the final model but was not significant. The multivariable model resulted in a relatively good prediction for the response of refusal, with an area under the ROC curve of 0.76 (Figure 2).

**Figure 2:**
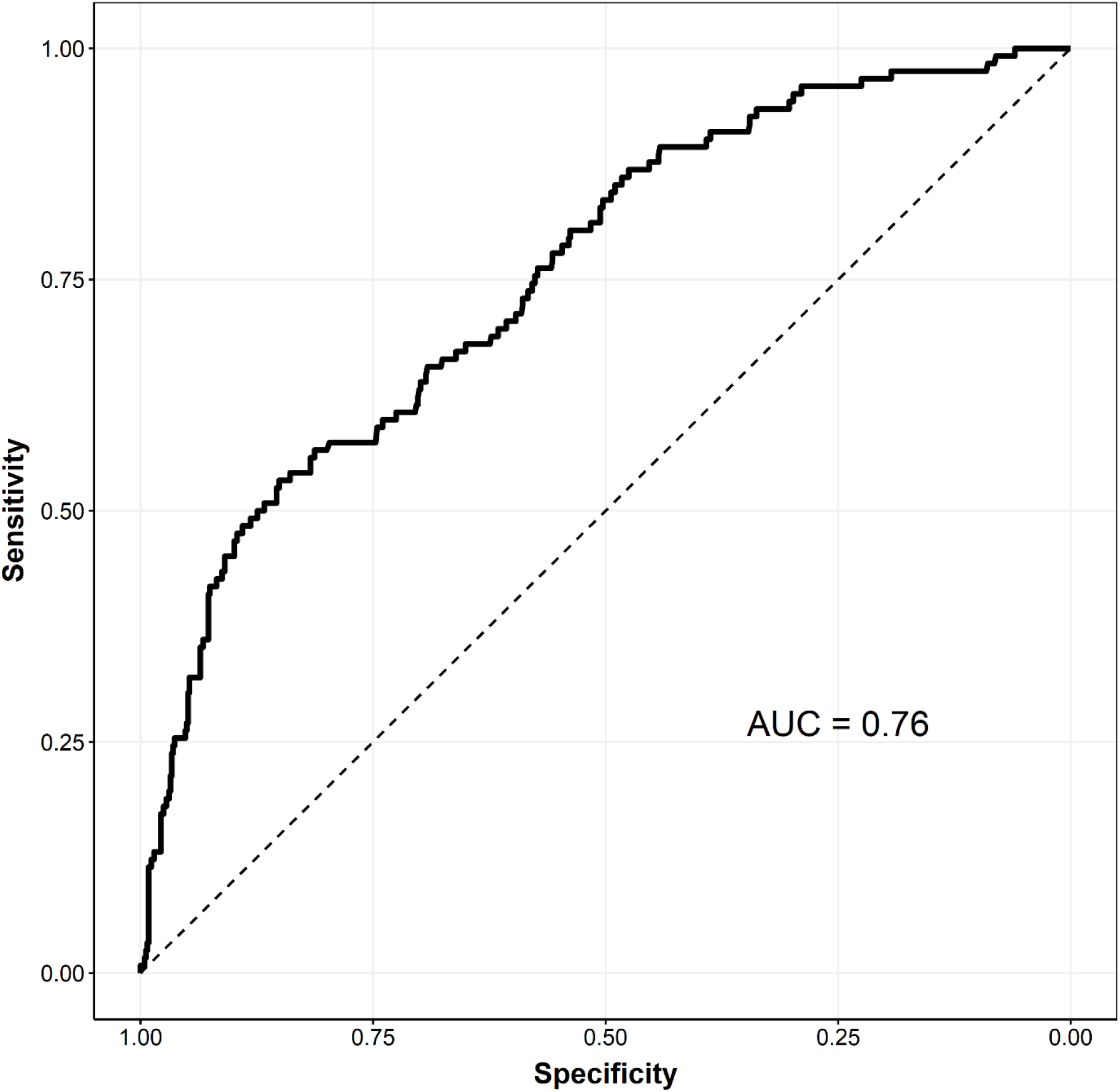
Receiver Operating Characteristics Curve for Prediction of Vaccine Refusal

## Discussion

Given the high morbidity and mortality from SARS-CoV-2 suffered by patients with dialysis dependent ESKD, it is important to understand vaccine acceptance in the “real world”. Dialysis providers, now with access to national vaccine stockpile, must plan rollouts that maximize acceptance, minimize waste and overcome hesitancy. Based on prior national surveys, we were surprised to find high vaccine acceptance rates in our dialysis program. Given the short time interval from notice of vaccine availability to mobile clinic launch, we had little time to provide formal vaccine education in advance. Providers and staff were encouraged to discuss vaccine benefits with patients, but no formal literature was provided aside from the day of the mobile vaccination clinic. This makes the primary outcome more surprising.

It is unclear which factors may have contributed to this (relatively) high acceptance rate. It is possible that the recent second surge in our geographic region with high infections among our dialysis population (∼ 16% of census) may have been a motivating factor. The vast majority of these infections occurred in the three months preceding the mobile vaccination clinic effort. The ability to provide vaccine access in-clinic clearly improved convenience. In addition, offering vaccine within patients’ “home” clinics by their care team may have reduced issues around vaccine trust that reduce acceptance (9). A peer effect was also possible – essentially undecided patients swayed to accept or refuse vaccination after observing peers vaccinated.

Curiously, we did not note a strong racial/ethnic difference in vaccine acceptance. Though black patients were modestly more likely to refuse vaccination than whites in univariate analysis, this did not rise to statistical significance, and there was no significant racial effect after multivariable adjustment. Extrapolating this finding to other national dialysis populations may be difficult given the much higher proportion of black patients in our program versus national estimates (10).

Patients that refused vaccine during the initial mobile vaccination clinics were more likely to be younger, female, and lack evidence of prior influenza vaccination. These findings are consistent with prior studies in the general public and dialysis patients (5,6,7). We hypothesized that prior infection with SARS-CoV-2 would reduce vaccine acceptance based on patient belief in natural immunity. This was not readily observed in our data. Of the 87 patients with prior infection history eligible for vaccine, 80% agreed to vaccination. Interestingly, the multivariable adjustment demonstrated increased vaccine refusal in association with longer dialysis vintage. The potential cause of this finding is unclear and we are not aware that this has been previously reported. Subsequent studies will be needed to further assess the relationship of both dialysis vintage and prior infection with SARS-CoV-2 to vaccine acceptance.

Operationally, several lessons were learned. Transportation problems endemic to dialysis patients necessitate in-center vaccination programs to achieve high acceptance rates. In addition, “catch-up” vaccination clinics for the 6% of patients not at their dialysis clinics during the mobile vaccination clinic and prior vaccine refusers changing their stance will be necessary to improve upon the attained 80% overall vaccination rate. This will require vaccine that is able to be safely stored and available on-demand at dialysis clinics. From a planning perspective, we have proposed a reasonably accurate predictive model of high likelihood vaccine refusers. Models such as these may be useful to identify and target patient populations at high risk of refusal.

Our study has several limitations. Given that the census of our program differs significantly from the national profile with a higher proportion of black patients and a lower census of latinx patients, some of our findings here may not directly translate to the national dialysis population. Also, unlike pre-vaccination surveys, we were not able to definitively obtain patient elements such as income and education level. Proxies for these were used, but may not be reliable. Additionally, due to limitations on data collection, unmeasured confounding factors may also have impacted our results. Finally, we were not able to directly verify receipt of the two-shot series in all patients obtaining vaccine outside of our mobile vaccine clinics (pharmacies, primary care offices, etc.). This may have caused underestimation of the proportion of patients fully vaccinated.

In summary, we present the results of an early dialysis program-wide vaccination effort. In our program, it appears that real-world vaccine acceptance is higher than early estimates in the general population and align with more recent surveys of the dialysis population. This data is encouraging. Patients at high risk of refusal share common attributes and likely can be identified for additional interventions to address vaccine hesitancy.

## Supporting information

Study Checklist

COI Lobo

COI Sharma

COI Bowman

COI Ma

## Data Availability

Data is not freely available externally. Please contact author if interested in de-identified data set for research purposes

## Acknowledgements

The authors gratefully acknowledge the assistance of the Virginia Department of Health / Blue Ridge Health district for providing vaccine access to UVA Dialysis patients and health care team members. The authors would also like to thank UVA Health pharmacists Lori Dunn PharmD and Elizabeth Stump PharmD, for their assistance and support with dialysis mobile vaccination clinics as well as Mr. Floyd Jennings for assistance with data acquisition. The authors would also like to all UVA Health dialysis vaccinators - especially our dedicated nursing staff.

